# Polygenic risk of idiopathic pulmonary fibrosis and COVID-19 severity

**DOI:** 10.1101/2023.06.12.23291269

**Authors:** Beatriz Guillen-Guio, Itahisa Marcelino-Rodriguez, Jose Miguel Lorenzo-Salazar, Olivia C Leavy, Richard J Allen, Scourge Cohort Group, José A. Riancho, Augusto Rojas, Pablo Lapunzina, Ángel Carracedo, Louise V Wain, Carlos Flores

## Abstract

**Introduction:** Coronavirus disease 2019 (COVID-19) survivors can develop residual lung abnormalities consistent with lung fibrosis. A shared genetic component between COVID-19 and idiopathic pulmonary fibrosis (IPF) has been shown. However, genetic overlap studies of IPF and COVID-19 have primarily concentrated on the IPF genome-wide significant risk variants that have been previously identified, rather than combined into a genome-wide polygenic risk. Here we used IPF genome-wide association study (GWAS) results to calculate polygenic risk scores (PRSs) and study their association with COVID-19 severity.

**Methods:** We used results from the largest meta-GWAS of clinically defined IPF risk (base dataset; n=24,589) and individual-level imputed data from the SCOURGE study of patients with COVID-19 (target dataset; n=15,024). We calculated IPF PRSs using PRSice-2 and assessed their association with COVID-19 hospitalisation, severe illness, and critical illness. We also evaluated the effect of age and sex stratification. Results were validated using an independent PRS method. Enrichment analyses and pathway-specific PRSs were performed to study biological pathways associated with COVID-19 severity.

**Results:** IPF PRSs were significantly associated with COVID-19 hospitalisation and severe illness. The strongest association was found in patients aged <60 years, especially among younger males (OR=1.16; 95%CI=1.08-1.25; p=6.39×10^−5^). A pathway enrichment analysis of the variants included in the best model fit and subsequent pathway-specific PRSs analyses supported the link of Cadherin and Integrin signalling pathways to COVID-19 severity when stratified by age and sex.

**Conclusion:** Our results suggest that there is genome-wide genetic overlap between IPF and severe COVID-19 that is dependent on age and sex and adds further support that the pathogenesis of both IPF and severe COVID-19 share underlying biological mechanisms. This could imply that individuals with a high IPF genetic risk are at an overall increased risk of developing lung sequelae resulting from severe COVID-19.

## Introduction

Coronavirus disease 2019 (COVID-19) is an infectious disease caused by the acute respiratory syndrome coronavirus 2 (SARS-CoV-2). Recent studies have shown that patients hospitalised by COVID-19 can develop residual lung abnormalities, including lung fibrosis [1, 2], although biological mechanisms underlying this process remain unclear. Idiopathic pulmonary fibrosis (IPF) is a chronic, progressive rare lung disease with a median survival of 3 years after diagnosis [3]. IPF has been shown to be an important risk factor for COVID-19 severity [4]. Additionally, studies have reported clinical and radiological similarities between severe COVID-19 and IPF [5]. Thus, there may be specific biological pathways that are common to the pathophysiology of both IPF and severe COVID-19.

Previous genetic studies have reported a shared causal genetic aetiology between IPF and COVID-19 severity [4, 6]. Out of the 19 common genetic variants previously reported for IPF risk, four of them were also associated with COVID-19 hospitalisation and severity, either increasing or decreasing disease risk, depending on the specific variant [6]. These include the most strongly associated IPF risk variant in genome-wide association studies (GWAS), located at the promoter of *MUC5B*, which displays an opposite direction of effect in IPF and COVID-19 (i.e., the IPF risk allele was protective for COVID-19). Interestingly, a mendelian randomisation (MR) analysis based on 15 genome-wide significant IPF risk variants known at the time revealed that, when the *MUC5B* locus was excluded, IPF had a causal effect on COVID-19 severity [4].

Prior studies assessing genetic overlap between IPF and COVID-19 have been based on sentinel variants associated with IPF susceptibility at the genome-wide significance level (p<5×10^−8^) in published GWAS. However, to date, no studies have evaluated the effect of combined whole-genome polygenic risk score (PRS) for IPF on COVID-19 severity. With the aim of identifying shared genes and biological pathways involved in the pathophysiology of both diseases, here we used a whole-genome PRS model to evaluate the genetic overlap between IPF and COVID-19 severity at genome-wide level and to assess if associations were age and/or sex dependent.

## Methods

### Study design and sample

We used IPF GWAS results (base dataset) to calculate PRSs for each individual in the COVID-19 dataset (target dataset) and tested the association of the PRSs with COVID-19 severity.

For IPF, we used publicly available summary statistic results from a large meta-GWAS of clinically defined IPF susceptibility comprising 4,125 IPF cases and 20,464 population controls of European ancestry from five different studies from UK, US, and Spain (https://github.com/genomicsITER/PFgenetics) [7]. All IPF cases were diagnosed according to the American Thoracic Society and European Respiratory Society guidelines [8, 9].

For COVID-19, we used individual-level genetic data (Single Nucleotide Polymorphism [SNP] arrays) from 11,939 individuals with a positive diagnosis of COVID-19 from the Spanish Coalition to Unlock Research on Host Genetics on COVID-19 (SCOURGE) study [10]. Patients were recruited between March and December 2020 and had not been vaccinated at the time of sample collection. Their data was collected and managed using REDCap electronic data capture tools hosted at Centro de Investigación Biomédica en Red (CIBER) [11, 12] (Supplemental Notes). We considered three previously defined COVID-19 phenotypes as cases to evaluate the response to COVID-19 infection: hospitalisation, severe illness (patients in scales 3 and 4), and critical illness (patients in scale 4), as per the severity scale defined by the SCOURGE consortium [10] (Table 1). We assigned as controls all other COVID-19 patients not satisfying the case condition, plus a total of 5,943 population controls from the Spanish DNA biobank (https://www.bancoadn.org) and GR@ACE consortium (Genome Research at Fundació ACE) [13]. Genotyping and quality control procedures have been previously described [10]. SNP Imputation was performed using the TOPMed version r2 reference panel (GRCh38) in the TOPMed Imputation Server [14, 15], and variants with a minor allele frequency (MAF)<1% or low imputation quality (Rsq<0.3) were filtered out. The SCOURGE study was performed in accordance with the Declaration of Helsinki and approved by the Galician Ethical Committee Ref 2020/197.

**Table 1.**
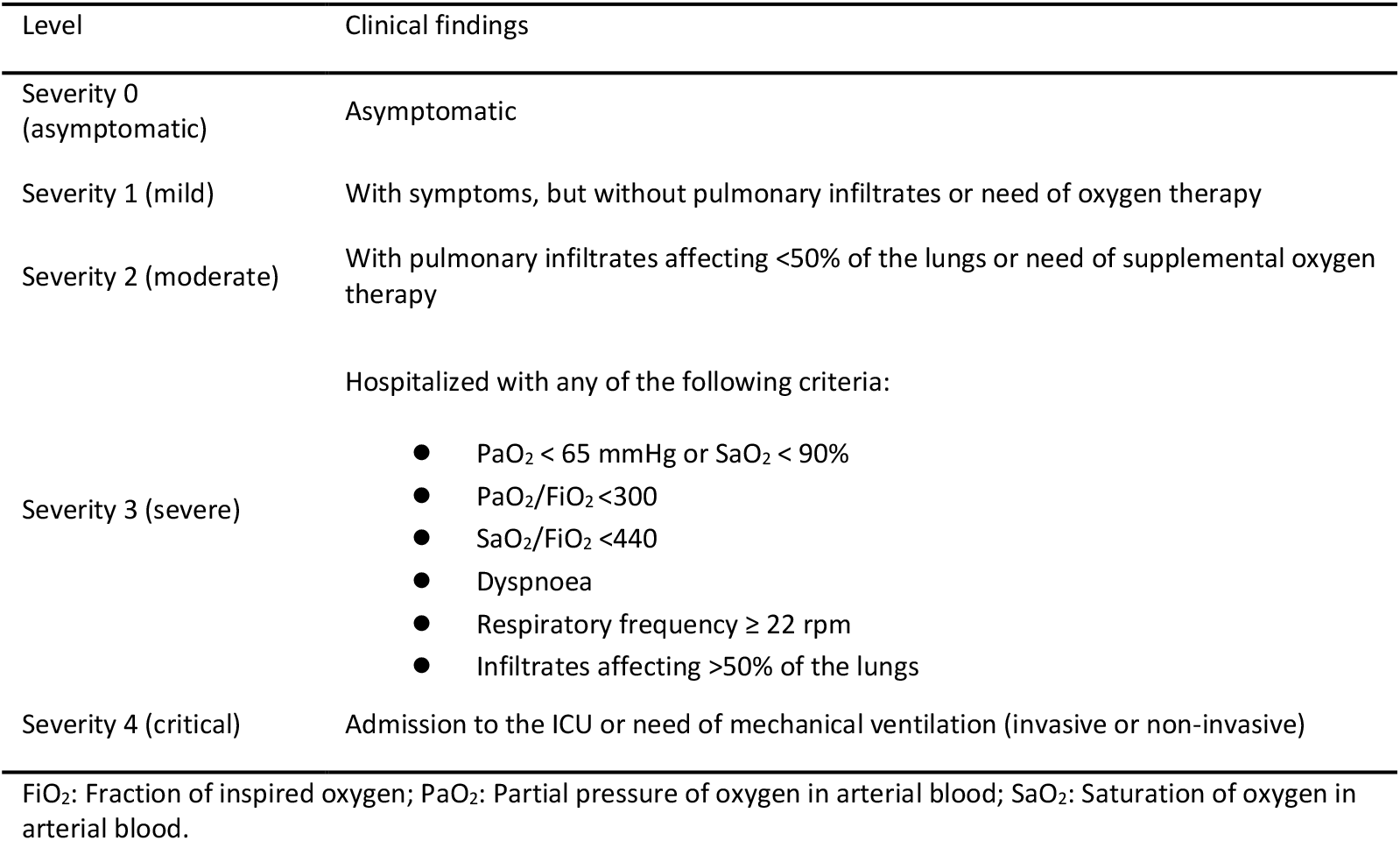
Classification of COVID-19 patients from SCOURGE into levels of severity.

Detailed information about both IPF and COVID-19 datasets can be found in the original publications describing the studies [7, 10].

### PRS modelling and statistical analyses

We used PRSice-2 [16] to calculate PRSs for each individual in the COVID-19 dataset. PRSs were estimated as the sum of risk alleles from the IPF GWAS variants weighted by their effect sizes:

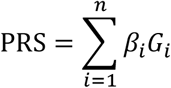

Where β is the weight or log Odds Ratio (OR) for variant *i, G* is the number of risk alleles carried at variant *i* and *n* is the number of variants included in the score. Briefly, PRSice-2 performs clumping to remove SNPs in high linkage disequilibrium (parameters were set at threshold R^2^= 0.1 for a 250 kb window for clumping), derives PRSs at different P-value thresholds in the base GWAS, tests the association of each of the PRS in the target dataset, and predicts the best model fit of the phenotype (i.e., the most significant model).

All PRSs were standardised as z-scores using the formula:

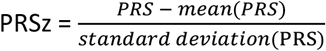

Binomial logistic regression was performed with R v4.0.3 [17] to assess the association of the individual PRS with COVID-19 hospitalisation, severe illness, and critical illness separately. To account for ascertainment bias, we considered a prevalence of COVID-19 hospitalisation of 0.5% according to previous estimates in the SCOURGE cohort [10]. Prevalence of severe illness (59% of hospitalised patients in the SCOURGE cohort) and critical illness (19% of hospitalisations) were estimated to be 0.295% and 0.095%, respectively [10]. We also performed stratifications by sex (male, female), age (<60 years, ≥60 years; to align with previous studies [4, 10, 18], and sex and age (male <60 years, male ≥60 years, female <60 years, female ≥60 years). All models were adjusted for the first 10 principal components (PC) for genetic ancestry and, whenever necessary, for age and sex. Significance was declared at p≤1.8×10^−3^ to control type-I error in the 27 comparisons (Bonferroni correction for nine models for three outcomes).

In addition, we performed the following as sensitivity analyses. To confirm the robustness of the PRS estimates in the significant comparisons, we calculated the individual PRSs using an alternative method with megaPRS [19], whose algorithm is based on variational Bayes. We also performed analyses excluding patients with a clinical history of chronic respiratory diseases or using only at-risk controls (i.e., non-hospitalised COVID-19 patients) as the control group. Additionally, we studied the predictive capacity of the PRSs on 90-day COVID-19 mortality (Bonferroni corrected p threshold=0.05/9=5.6×10^−3^). Finally, we compared the effect of whole-genome PRS calculated from genome-wide variants with a PRS obtained from only the 19 previously published genome-wide significant IPF common risk variants (sentinels PRS) (**Table S1**) [7]. These comparisons were also tested excluding the *MUC5B* locus, the strongest genetic risk factor for IPF, either by excluding the *MUC5B* promoter variant (**Table S1**) from the sentinels PRS model or by excluding all the variants in the ±1 Mb flanking regions from the whole-genome PRS model.

### Pathway-specific PRS analyses

We first used the online data tool GREAT [20] to obtain the list of genes that were experimentally or computationally linked through regulatory annotations to the genetic variants included in the best PRS model of COVID-19 hospitalisation for the entire study sample (i.e., the variants selected by PRSice-2 for providing the Best P-value Threshold, **Table 2**). We then performed a gene set enrichment analysis with ShinyGO 0.77 [21] to assess if the obtained gene list was enriched in genes involved in certain biological pathways (False Discovery Rate (FDR)< 0.05). For the top five pathways overrepresented in the gene list, we calculated pathway-specific PRSs by including only the variants associated with each of the five pathways separately in the PRS computation, and then studied their association with COVID-19 hospitalisation. Significance was declared at p<0.05.

**Table 2.** Association of IPF PRS models with COVID-19 severity.

| COVID-19 severity | Category | Covariates | Cases/Controls* | PT | Num_SNP | OR (95%CI) | P-Value† |
| --- | --- | --- | --- | --- | --- | --- | --- |
| Hospitalisation | All individuals | Age, Sex, 10PC | 5,968/9056 | 2.10x10<sup>-3</sup> | 2,939 | 1.08<br>(1.04,1.12) | 7.90x10<sup>-5</sup> |
|  | Males | Age, 10PC | 3,441/3,958 | 7.85x10<sup>-3</sup> | 8,686 | 1.09<br>(1.04,1.16) | 1.28x10<sup>-3</sup> |
|  | Females | Age, 10PC | 2,525/5,096 | 2.25x10<sup>-3</sup> | 3,083 | 1.08<br>(1.02,1.14) | 6.44x10<sup>-3</sup> |
|  | ≥60 years | Sex, 10PC | 4,328/2,355 | 7.50x10<sup>-4</sup> | 1,330 | 1.06<br>(1.01,1.12) | 0.026 |
|  | <60 years | Sex, 10PC | 1,607/6,457 | 4.70x10<sup>-3</sup> | 5,660 | 1.12<br>(1.06,1.19) | 3.44x10<sup>-5</sup> |
|  | ≥60 years Males | 10PC | 2,436/878 | 7.85x10<sup>-3</sup> | 8,686 | 1.06<br>(0.98,1.15) | 0.123 |
|  | ≥60 years Females | 10PC | 1,892/1,477 | 7.50x10<sup>-4</sup> | 1,330 | 1.09<br>(1.01,1.17) | 0.021 |
|  | <60 years Males | 10PC | 987/2,904 | 4.70x10<sup>-3</sup> | 5,660 | 1.16<br>(1.08,1.25) | 6.39x10<sup>-5</sup> |
|  | <60 years Females | 10PC | 619/3,551 | 0.070 | 51,746 | 1.13<br>(1.04,1.23) | 6.04x10<sup>-3</sup> |
| Severe illness | All individuals | Age, Sex, 10PC | 3,502/10,846 | 6.65x10<sup>-3</sup> | 7,566 | 1.08<br>(1.04,1.13) | 2.57x10<sup>-4</sup> |
|  | Males | Age, 10PC | 2,109/4,897 | 6.40x10<sup>-3</sup> | 7,331 | 1.08<br>(1.02,1.14) | 6.05x10<sup>-3</sup> |
|  | Females | Age, 10PC | 1,393/5,949 | 0.014 | 13,949 | 1.09<br>(1.02,1.16) | 7.93x10<sup>-3</sup> |
|  | ≥60 years | Sex, 10PC | 2,543/3,834 | 0.014 | 14,289 | 1.05<br>(1.00,1.11) | 0.057 |
|  | <60 years | Sex, 10PC | 959/7,012 | 3.30x10<sup>-3</sup> | 4,198 | 1.14<br>(1.07,1.22) | 1.53x10<sup>-4</sup> |
|  | ≥60 years Males | 10PC | 1,526/1,631 | 1.00 | 308,260 | 1.06<br>(0.99,1.14) | 0.095 |
|  | ≥60 years Females | 10PC | 1,017/2,203 | 0.014 | 13,949 | 1.08<br>(1.00,1.17) | 0.047 |
|  | <60 years Males | 10PC | 583/3,266 | 5.05x10<sup>-3</sup> | 6,009 | 1.16<br>(1.06,1.27) | 9.98x10<sup>-4</sup> |
|  | <60 years Females | 10PC | 376/3,746 | 0.073 | 53,482 | 1.15<br>(1.03,1.28) | 0.010 |
| Critical illness | All individuals | Age, Sex, 10PC | 1,124/13,224 | 0.014 | 13,828 | 1.12<br>(1.06,1.20) | 2.49x10<sup>-4</sup> |
|  | Males | Age, 10PC | 815/6,191 | 1.90x10<sup>-3</sup> | 2,728 | 1.12<br>(1.04,1.21) | 2.10x10<sup>-3</sup> |
|  | Females | Age, 10PC | 309/7,033 | 0.053 | 41,838 | 1.23<br>(1.10,1.39) | 3.67x10<sup>-4</sup> |
|  | ≥60 years | Sex, 10PC | 776/5,601 | 0.014 | 13,949 | 1.13<br>(1.04,1.21) | 2.36x10<sup>-3</sup> |
|  | <60 years | Sex, 10PC | 348/7,623 | 0.047 | 37,678 | 1.16<br>(1.04,1.30) | 6.83x10<sup>-3</sup> |
|  | ≥60 years Males | 10PC | 554/2,603 | 3.10x10<sup>-3</sup> | 3,961 | 1.15<br>(1.05,1.26) | 2.92x10<sup>-3</sup> |
|  | ≥60 years Females | 10PC | 222/2,998 | 0.053 | 41,863 | 1.18<br>(1.03,1.36) | 0.017 |
|  | <60 years Males | 10PC | 261/3,588 | 7.45x10<sup>-3</sup> | 8,328 | 1.11<br>(0.98,1.26) | 0.090 |
|  | <60 years Females | 10PC | 87/4,035 | 0.129 | 84,188 | 1.44<br>(1.16,1.78) | 8.97x10<sup>-4</sup> |

\*A total of 281 individuals had missing data for sex (n=4) and age (n=277).
†Association results for the best prediction model resulting from PRSice-2 in each category (the PRS includes all variants reaching the best P-value threshold in the IPF GWAS). Significance declared at $p \leq 1.85 \times 10^{-3}$ after Bonferroni correction.
PC: Principal Components, PT: Best fit P-value Threshold; Num\_SNP: Number of SNPs included in the best model.

## Results

The COVID-19 SCOURGE dataset comprised a total of 15,024 individuals of European genetic ancestry. Sample sizes for each COVID-19 phenotype and stratification category are summarised in **Table 2**. A total of 308,260 clumped variants from the IPF meta-GWAS were used for PRS analyses with PRSice-2.

IPF whole-genome PRS were found to be associated with COVID-19 hospitalisation (OR= 1.08; 95% Confidence Interval [CI]=1.04-1.12; p=7.90×10^−5^), severe illness (OR=1.08; 95% CI=1.04-1.13; p=2.57×10^−4^), and critical illness (OR=1.12; 95% CI=1.06-1.20; p=2.49×10^−4^). Stratifying by age and/or sex, the strongest association was found in hospitalised younger individuals (<60 years), especially among younger males (**Table 2**). Among these categories, the lowest number of IPF genetic variants that were needed to calculate the best performing whole-genome PRS was 2,939 variants, all of them reaching a p-value threshold of 2.10×10^−3^ in the GWAS of IPF (**Table 2, Figure S1**). A validation of the significant PRSs associations based on megaPRS estimations supported the robustness of associations of the IPF whole-genome PRS with COVID-19 hospitalisation and severe illness (**Table S2**). The validation results for critical illness were inconsistent, potentially attributed to the limited sample size available for this particular phenotype.

Post-hoc analyses excluding 892 patients with a clinical history of chronic respiratory diseases supported the robustness of the results in the significant hospitalisation categories (**Table S3**). When comparing hospitalised COVID-19 patients with individuals with COVID-19 who were not hospitalised, association results were consistent across all categories (**Table S4**). IPF whole-genome PRSs did not show an association with 90-day COVID-19 mortality in the SCOURGE study for any of the best models in each category (**Table S5**).

The exclusion of the *MUC5B* locus (promoter variant and variants in the ±1 Mb flanking regions) from the IPF PRS modelling resulted in a stronger association of the IPF whole-genome PRS with COVID-19 hospitalisation (**Table S6**). When using sentinels PRSs calculated from the previously known IPF risk variants, the association with COVID-19 hospitalisation was only significant for models where the *MUC5B* locus had been excluded (**Table S6**). We next compared the results of the IPF whole-genome PRS to those of the sentinels PRS excluding the *MUC5B* locus. When considering all individuals or all younger (<60 years old) individuals, the effect for COVID-19 hospitalisation was larger when whole-genome PRSs were used (**Table S6**). However, among males, the effect size was bigger when excluding *MUC5B* from the sentinels PRS. The effect didn’t differ in <60 years old males.

A gene set enrichment analysis was performed to identify the biological processes that may be related to both IPF and COVID-19 hospitalisation. We focused on the 3,471 genes linked through experimental and computational regulatory annotations by GREAT to the 2,939 SNPs included in the whole-genome PRS model associated with COVID-19 hospitalisation in the whole sample (**Table 2**). Results showed a significant enrichment in genes involved in Cadherin, Wnt and Integrin signalling pathways, among others (**Figure S2A**). Results were similar when the 19 IPF common loci (sentinels ±1MB) were excluded from the analysis (**Figure S2B**) and when we used the list of 5,660 SNPs used to calculate the best-fit PRS associated with COVID-19 hospitalisation in <60 years males (**Table 2, Figure S3**). Pathway-specific PRSs analyses revealed that the PRS including the Integrin pathway variants was significantly associated with COVID-19 hospitalisation in patients younger than 60 years (p=7.0×10^−3^) (**Table S7**). Additionally, the Cadherin pathway PRS was associated with COVID-19 hospitalisation in males (p=0.028) (**Table S7**). No significant associations were observed for the remaining pathways.

## Discussion

We performed a PRS-based association analysis to study the genetic overlap between IPF and the severity of COVID-19. Our results show that IPF PRSs obtained from genome-wide variants were significantly associated with COVID-19 hospitalisation and severe illness, suggesting that IPF and severe COVID-19 could share certain biological mechanisms. Our results were also robust when using an alternate method to estimate the PRSs, excluding patients with previously reported chronic respiratory diseases and using non-hospitalised COVID-19 patients as controls (instead of relying on population-based controls with uncertain SARS-CoV-2 exposure at the time of data collection).

The most robust associations of IPF PRS models with COVID-19 were found for the hospitalised patients who were younger (<60 years old), particularly in younger males. These results agree with Fadista and colleagues’ findings, who showed a modest protective effect of the IPF risk-allele at *MUC5B* for COVID-19 hospitalisation in >60 years old individuals [4]. Additionally, Nakanishi and colleagues [22] reported that the effects of carrying common genetic risk factors of severe COVID-19 were stronger in individuals younger than 60 years. Cruz and colleagues also showed that the genetic risk score combining the main COVID-19 genetic risk factors had a higher predictive capability among the <60 years old males from the SCOURGE study [10]. Taken together, this reinforces the existence of biological mechanisms shared between IPF and severe COVID-19 pathogenesis and supports that the presence of genetic risk factors that affect disease severity would be more evident among younger patients, while severe COVID-19 in older individuals (≥60 years old) might be more influenced by non-genetic factors such as comorbidities and immunological defects [23].

The existence of genetic factors shared between IPF and COVID-19 had already been described by several studies [4, 6]. However, here we suggest that the number of overlapping variants underlying IPF risk and severe COVID-19 could be larger than previously reported. In fact, the sentinels PRS calculated based on only the previously reported IPF risk variants was not significantly associated with severe COVID-19 in our data. However, when the strongest known IPF genetic risk factor at *MUC5B* was excluded from the sentinels PRS model, the effect on COVID-19 hospitalisation was stronger, as previously reported by Fadista and colleagues [4]. This could be explained by the different direction of effect that the *MUC5B* variant has for IPF and COVID-19 [4, 6]. After excluding the *MUC5B* locus, the effect for COVID-19 hospitalisation was larger in all and <60 years old patients when using whole-genome PRSs compared to sentinels PRSs results. Nevertheless, the effect on COVID-19 hospitalisation was similar with both approaches in males and younger males. This could also be attributed to the arbitrary linkage disequilibrium clumping method that was used to select variants to be included in the whole-genome PRS models, compared to the refined selection of the 19 IPF risk variants involved in the sentinels PRS models.

Our gene set enrichment and pathway-specific PRS analyses suggest that Integrin and Cadherin signalling pathways, previously related to fibrosis and lung repair processes, could have a role in COVID-19 severity. These could be of special interest among young (<60 years) patients and males based on our results. Integrins are considered key regulators during fibrogenesis, and integrin inhibitors are currently being investigated as anti-fibrotic strategies for IPF treatment, including and inhaled inhibitor of the αvβ6 integrin [24, 25]. Additionally, several studies have reported the role of integrins in facilitating the cellular entry of SARS-CoV-2 [26, 27]. Therefore, this pathway could have significant potential as a key target for the treatment of both SARS-CoV-2 infection and post-COVID-19 pulmonary fibrosis. Signalling through cadherins has also been suggested to be involved in lung fibrosis [28]. Cadherin-11 may play an important role in the pathogenesis of IPF, possibly through the regulation of epithelial to mesenchymal transition (EMT) in alveolar epithelial cells [29], again pointing to this protein activity as a potential therapeutic target. Recent studies suggest that the EMT process could also be key in post-COVID-19 lung fibrosis [30].

Despite the evidence supporting the genetic overlap of IPF risk and severe COVID-19 [4, 6], the main limitations of the study are: i) the lack of an independent cohort study to validate the association of IPF PRS with severe COVID-19; and ii) the lack of genetic diversity in SCOURGE since it is composed primarily of European genetic ancestry patients. This makes it difficult to anticipate the generalisability of findings to populations of other genetic ancestries. Among the strengths, it is worth highlighting that COVID-19 participants of SCOURGE were recruited before vaccines were widely available for the community, reducing potential biases caused by disease severity misclassification. Furthermore, contrary to previous studies, the whole-genome PRS approach enables inclusion of genetic variants associated with IPF that have not yet been reported due to limitations of statistical power and strict significance thresholds necessitated by multiple testing corrections.

In summary, the use of a whole-genome PRS approach supported the existence of thousands of shared common genetic risk factors underlying the pathogenesis of both IPF and severe COVID-19. This comprehensive approach was more effective in capturing the severity of COVID-19 compared to an IPF-sentinels only approach. According to our results, overlapping variants could be involved in biological processes such as Integrin and Cadherin signalling pathways. Furthermore, we observed that the association of the polygenic risks of IPF with COVID-19 hospitalisation was age- and sex-dependent. Given that an increased genetic risk for IPF is associated with a higher risk of severe COVID-19, and that severe COVID-19 also increases risk of post COVID-19 lung sequelae among survivors [2], individuals at high IPF genetic risk could be at an overall increased risk of developing post-COVID-19 lung fibrotic sequelae. This could provide valuable insights for shaping future therapeutic strategies in managing COVID-19. Further studies will be needed to evaluate this possibility.

## Data Availability

All data produced in the present study are available upon reasonable request to the authors

## Authors contribution

BG-G, IM-R, JML-S, OCL, and RJA performed the analyses. JAR, AR, PL, AC, LVW, and CF participated in data collection. LVW and CF supervised the study. BG-G, AC, LVW, and CF obtained funding. BG-G and CF wrote the first draft of the manuscript. All authors revised and approved the final version.

