## Supplement for "Polygenic risk of idiopathic pulmonary fibrosis and COVID-19 severity"

#### Supplementary material

Beatriz Guillen-Guio*, Itahisa Marcelino-Rodriguez, Jose Miguel Lorenzo-Salazar, Olivia C Leavy, Richard J Allen, Scourge Cohort Group, José A. Riancho, Augusto Rojas, Pablo Lapunzina, Ángel Carracedo, Louise V Wain#, Carlos Flores#

#Equal contribution as senior authors

### Supplemental Note

**Research electronic data capture (REDCap)** is a secure, web-based software platform designed to support data capture for research studies, providing 1) an intuitive interface for validated data capture; 2) audit trails for tracking data manipulation and export procedures; 3) automated export procedures for seamless data downloads to common statistical packages; and 4) procedures for data integration and interoperability with external sources [1, 2].

### Supplementary Tables

**Table S1. IPF sentinel variants**

| Locus | SNP ID | Chr. | Position (hg38) | Effect allele | Non-effect allele | OR [95% CI] | P-value[f] |
|---|---|---|---|---|---|---|---|
| *KIF15* | rs141979279[a] | 3 | 44816639 | C | T | 1.50 [1.33,1.70] | $1.21x10^{-10}$ |
| *TERC* | rs10936601[b] | 3 | 169810661 | C | T | 0.79 [0.74,0.84] | $2.10x10^{-15}$ |
| *FAM13A* | rs2013701 | 4 | 88963935 | G | T | 1.25 [1.18,1.32] | $4.60x10^{-16}$ |
| *TERT* | rs7725218 | 5 | 1282299 | G | A | 1.41 [1.33,1.50] | $4.90x10^{-32}$ |
| *DSP* | rs2076295 | 6 | 7562999 | G | T | 1.49 [1.41,1.57] | $1.50x10^{-48}$ |
| *MAD1L1* | rs12699415 | 7 | 1869843 | A | G | 1.27 [1.20,1.34] | $7.85x10^{-18}$ |
| *ZKSCAN1* | rs2897075 | 7 | 100032719 | T | C | 1.30 [1.23,1.37] | $1.77x10^{-21}$ |
| *DEPTOR* | rs28513081 | 8 | 119921886 | A | G | 1.20 [1.13,1.27] | $1.22x10^{-9}$ |
| *10q25.1* | rs79684490 | 10 | 109470103 | A | G | 1.40 [1.24,1.57] | $3.52x10^{-8}$ |
| *MUC5B* | rs35705950 | 11 | 1219991 | T | G | 5.06 [4.69,5.47] | $9.09x10^{-418}$ |
| *ATP11A* | rs12585036[c] | 13 | 112881427 | C | T | 1.29 [1.21,1.38] | $5.99x10^{-14}$ |
| *IVD* | rs59424629 | 15 | 40428343 | T | G | 1.27 [1.21,1.34] | $4.98x10^{-19}$ |
| *KNL1* | rs12912339 | 15 | 40639510 | A | G | 1.30 [1.21,1.39] | $7.41x10^{-13}$ |
| *AKAP13* | rs62023891 | 15 | 85553985 | A | G | 1.18 [1.12,1.25] | $1.32x10^{-8}$ |
| *NPRL3* | rs74614704 | 16 | 112241 | A | G | 1.49 [1.34,1.67] | $2.57x10^{-12}$ |
| *17q21.31* | rs3785884[d] | 17 | 45980229 | G | A | 1.40 [1.30,1.51] | $2.53x10^{-20}$ |
| *DPP9* | rs35574495[e] | 19 | 4686976 | G | T | 0.80 [0.75,0.86] | $1.08x10^{-9}$ |
| *STMN3* | rs112087793 | 20 | 63652817 | C | T | 1.34 [1.21,1.48] | $1.09x10^{-8}$ |
| *RTEL1* | rs41308092 | 20 | 63693038 | A | G | 1.75 [1.45,2.10] | $3.13x10^{-9}$ |

[a]Proxy for rs78238620 ($r^2$=1); [b]Proxy for rs12696304 ($r^2$=0.99); [c]Proxy for rs9577395 ($r^2$=0.98); [d]Proxy for rs2077551 ($r^2$=0.83); [e]Proxy for rs12610495 ($r^2$=0.36); [f]Significance of the association in the GWAS of IPF susceptibility [3]. Linkage disequilibrium ($r^2$) proxies were defined using 1000 genomes European sample data. Data for the *SPDL1* locus in SCOURGE was no available given the rarity (<1%) of the IPF risk variant in the general population [4].

**Table S2. Validation of IPF PRS associations with COVID-19 severity using an alternative PRS estimation method**

| COVID-19 severity | Category | PRSice-2* | | | | megaPRS | |
|---|---|---|---|---|---|---|---|
| | | PT | Num_SNP | OR (95%CI) | P-Value | OR (95%CI) | P-Value |
| Hospitalisation | All individuals | $2.10x10^{-3}$ | 2,939 | 1.08 (1.04,1.12) | $7.90x10^{-5}$ | 1.07 (1.03,1.11) | $1.27x10^{-3}$ |
| | Males | $7.85x10^{-3}$ | 8,686 | 1.09 (1.04,1.16) | $1.28x10^{-3}$ | 1.08 (1.02,1.14) | $9.10x10^{-3}$ |
| | <60 years | $4.70x10^{-3}$ | 5,660 | 1.12 (1.06,1.19) | $3.44x10^{-5}$ | 1.10 (1.04,1.16) | $1.17x10^{-3}$ |
| | <60 years Males | $4.70x10^{-3}$ | 5,660 | 1.16 (1.08,1.25) | $6.39x10^{-5}$ | 1.13 (1.05,1.21) | $1.55x10^{-3}$ |
| Severe illness | All individuals | $6.65x10^{-3}$ | 7,566 | 1.08 (1.04,1.13) | $2.57x10^{-4}$ | 1.06 (1.02,1.11) | $4.80x10^{-3}$ |
| | <60 years | $3.30x10^{-3}$ | 4,198 | 1.14 (1.07,1.22) | $1.53x10^{-4}$ | 1.14 (1.06,1.22) | $1.79x10^{-4}$ |
| | <60 years Males | $5.05x10^{-3}$ | 6,009 | 1.16 (1.06,1.27) | $9.98x10^{-4}$ | 1.18 (1.08,1.29) | $2.02x10^{-4}$ |
| Critical illness | All individuals | 0.014 | 13,828 | 1.12 (1.06,1.20) | $2.49x10^{-4}$ | 1.08 (1.01,1.15) | 0.017 |
| | Females | 0.053 | 41,838 | 1.23 (1.10,1.39) | $3.67x10^{-4}$ | 1.14 (1.01,1.27) | 0.029 |
| | <60 years Females | 0.129 | 84,188 | 1.44 (1.16,1.78) | $8.97x10^{-4}$ | 1.09 (0.88,1.34) | 0.426 |

*Association results for the best prediction model resulting from PRSice-2 in significant categories ($p \leq 1.85x10^{-3}$). The PRSice-2 PRS includes all variants reaching the best P-value threshold in the IPF GWAS.
PC: Principal Components, PT: Best fit P-value Threshold; Num_SNP: Number of SNPs included in the best model.

**Table S3. Association of IPF whole-genome PRS with COVID-19 hospitalisation in the entire sample *vs.* after excluding patients with clinical history of chronic respiratory diseases**

| COVID-19 hospitalisation category* | All individuals (Nt=15,024) | | | Excluding chronic diseases (Nt=14,132) † | | |
|---|---|---|---|---|---|---|
| | PT | OR (95% CI) | P-value | PT | OR (95% CI) | P-value |
| All | $2.10x10^{-3}$ | 1.08 (1.04,1.12) | $7.90x10^{-5}$ | $2.30x10^{-3}$ | 1.09 (1.04,1.13) | $4.31x10^{-5}$ |
| Males | $7.85x10^{-3}$ | 1.09 (1.04,1.16) | $1.28x10^{-3}$ | $7.85x10^{-3}$ | 1.10 (1.04,1.17) | $7.89x10^{-4}$ |
| <60 years | $4.70x10^{-3}$ | 1.12 (1.06,1.19) | $3.44x10^{-5}$ | $4.70x10^{-3}$ | 1.13 (1.07,1.20) | $2.62x10^{-5}$ |
| <60 years Males | $4.70x10^{-3}$ | 1.16 (1.08,1.25) | $6.39x10^{-5}$ | $4.70x10^{-3}$ | 1.17 (1.08,1.26) | $7.08x10^{-5}$ |

Association results for the best prediction model resulting from PRSice-2. Nt: total sample size; PT: Best fit P-value Threshold.
†A total of 892 patients with chronic respiratory diseases were excluded

**Table S4. Association of IPF whole-genome PRS with COVID-19 hospitalisation including or excluding population controls**

| COVID-19 hospitalisation category | Non-hospitalised COVID-19 + population controls | | | | Non-hospitalised COVID-19 controls | | | |
|---|---|---|---|---|---|---|---|---|
| | PT | Cases/Controls* | OR (95% CI) | P-value | PT | Cases/Controls* | OR (95% CI) | P-value |
| All | $2.10 \times 10^{-3}$ | 5,968/9,056 | 1.08 (1.04,1.12) | $7.90 \times 10^{-5}$ | $2.30 \times 10^{-3}$ | 5,968/3,382 | 1.06 (1.01,1.12) | 0.020 |
| Males | $7.85 \times 10^{-3}$ | 3,441/3,958 | 1.09 (1.04,1.16) | $1.28 \times 10^{-3}$ | 0.049 | 3,441/912 | 1.07 (1.01,1.13) | 0.114 |
| <60 years | $4.70 \times 10^{-3}$ | 1,607/6,457 | 1.12 (1.06,1.19) | $3.44 \times 10^{-5}$ | 0.069 | 1,607/2,567 | 1.12 (1.05,1.20) | $1.07 \times 10^{-3}$ |
| <60 years Males | $4.70 \times 10^{-3}$ | 987/2,904 | 1.16 (1.08,1.25) | $6.39 \times 10^{-5}$ | $7.85 \times 10^{-3}$ | 987/649 | 1.12 (1.02,1.24) | 0.020 |

Association results for the best prediction model resulting from PRSice-2. PT: Best fit P-value Threshold. *A total of 281 individuals had missing data for sex (n=4) and age (n=277).

**Table S5. Association of IPF whole-genome PRS with 90-day COVID-19 mortality**

| COVID-19 mortality category | Covariates | All individuals (Nt: 758 cases and 7,508 controls) | | | Excluding chronic respiratory diseases (Nt: 649 cases and 6,797 controls)† | | |
|---|---|---|---|---|---|---|---|
| | | PT | OR (95% CI) | P-value | PT | OR (95% CI) | P-value |
| All | Age, Sex, 10PC | $1.70 \times 10^{-3}$ | 1.11 (1.02,1.20) | 0.012 | $1.70 \times 10^{-3}$ | 1.11 (1.01,1.21) | 0.023 |
| Males | Age, 10PC | $8.5 \times 10^{-4}$ | 1.14 (1.03,1.27) | 0.014 | $1.00 \times 10^{-4}$ | 1.16 (1.03,1.31) | 0.012 |
| Females | Age, 10PC | $5.0 \times 10^{-4}$ | 1.10 (0.96,1.25) | 0.157 | $4.3 \times 10^{-3}$ | 1.07 (0.93,1.22) | 0.336 |
| ≥60 years | Sex, 10PC | $9.5 \times 10^{-4}$ | 1.09 (1.01,1.18) | 0.033 | $9.5 \times 10^{-4}$ | 1.08 (0.99,1.18) | 0.081 |
| <60 years | Sex, 10PC | $4.5 \times 10^{-4}$ | 1.27 (0.99,1.65) | 0.063 | 0.011 | 1.27 (0.97,1.66) | 0.083 |
| ≥60 years Males | 10PC | $1.00 \times 10^{-3}$ | 1.13 (1.01,1.26) | 0.028 | $1.00 \times 10^{-4}$ | 1.16 (1.02,1.30) | 0.019 |
| ≥60 years Females | 10PC | 0.416 | 0.91 (0.80,1.03) | 0.149 | 0.351 | 0.90 (0.79,1.03) | 0.121 |
| <60 years Males | 10PC | 0.460 | 0.75 (0.53,1.06) | 0.108 | 0.460 | 0.74 (0.51,1.06) | 0.097 |
| <60 years Females | 10PC | 0.011 | 1.55 (1.03,2.34) | 0.035 | 0.011 | 1.64 (1.07,2.53) | 0.024 |

PRSice-2 association results for the best prediction model in each category. †A total of 820 patients with chronic respiratory diseases were excluded. Nt: total sample size; PC: Principal Components; PT: Best fit P-value Threshold. Validation significance declared at $p<5.6 \times 10^{-3}$ after Bonferroni correction (0.05/9 tests).

#### Table S6. Association with COVID-19 hospitalisation after excluding the *MUC5B* locus

| | | | Whole-genome PRS | | | Sentinels PRS† | |
|---|---|---|---|---|---|---|---|
| COVID-19 hospitalisation category | Covariates | *MUC5B* included | PT | OR (95% CI) | P-value* | OR (95%CI) | P-value |
| All | Age, Sex, 10PC | Yes | $2.10 \times 10^{-3}$ | 1.08 (1.04,1.12) | $7.90 \times 10^{-5}$ | 1.01 (0.97,1.05) | 0.772 |
| | | No | $2.10 \times 10^{-3}$ | 1.09 (1.05,1.14) | $5.55 \times 10^{-6}$ | 1.06 (1.02,1.10) | 0.005 |
| Males | Age, 10PC | Yes | $7.85 \times 10^{-3}$ | 1.09 (1.04,1.16) | $1.28 \times 10^{-3}$ | 1.05 (0.99,1.10) | 0.107 |
| | | No | $7.85 \times 10^{-3}$ | 1.10 (1.04,1.17) | $4.39 \times 10^{-4}$ | 1.12 (1.06,1.18) | $7.55 \times 10^{-5}$ |
| <60 years | Sex, 10PC | Yes | $4.70 \times 10^{-3}$ | 1.12 (1.06,1.19) | $3.44 \times 10^{-5}$ | 1.02 (0.97,1.08) | 0.415 |
| | | No | $4.70 \times 10^{-3}$ | 1.14 (1.08,1.21) | $3.11 \times 10^{-6}$ | 1.10 (1.04,1.16) | $9.34 \times 10^{-4}$ |
| <60 years Males | 10PC | Yes | $4.70 \times 10^{-3}$ | 1.16 (1.08,1.25) | $6.39 \times 10^{-5}$ | 1.08 (1.00,1.16) | 0.039 |
| | | No | $4.70 \times 10^{-3}$ | 1.19 (1.10,1.28) | $5.56 \times 10^{-6}$ | 1.19 (1.10,1.28) | $3.67 \times 10^{-6}$ |

To assess the PRS models after excluding *MUC5B*, the *MUC5B* promoter variant rs35705950 and variants within ±1 Mb were removed from the analyses. PC: Principal Components; PT: Best fit P-value Threshold. *PRSice-2 association results for the best prediction model in each category. †Association results for PRS calculated using the 19 previously reported genome-wide significant IPF common risk variants shown in Table S1. No thresholding was applied to sentinels PRS analyses.

#### Table S7. Results from the pathway specific PRS analyses

| Pathway [Number of SNPs included in the PRS*] | COVID-19 hospitalisation category | Co-variables | OR (95%CI) | P-value |
|---|---|---|---|---|
| Cadherin pathway [69] | All | Age, Sex, 10PC | 1.03 (0.99, 1.07) | 0.171 |
| | Males | Age, 10PC | 1.06 (1.01, 1.12) | **0.028** |
| | <60 years | Sex, 10PC | 1.03 (0.98, 1.09) | 0.249 |
| | <60 years Males | 10PC | 1.07 (1.00, 1.15) | 0.057 |
| Wnt pathway [102] | All | Age, Sex, 10PC | 1.01 (0.97, 1.05) | 0.531 |
| | Males | Age, 10PC | 1.02 (0.96, 1.07) | 0.584 |
| | <60 years | Sex, 10PC | 1.02 (0.96, 1.08) | 0.524 |
| | <60 years Males | 10PC | 1.02 (0.95, 1.09) | 0.649 |
| Integrin pathway [73] | All | Age, Sex, 10PC | 1.03 (0.99, 1.07) | 0.103 |
| | Males | Age, 10PC | 1.03 (0.97, 1.08) | 0.352 |
| | <60 years | Sex, 10PC | 1.08 (1.02, 1.14) | **0.007** |
| | <60 years Males | 10PC | 1.07 (0.99, 1.15) | 0.078 |
| Heterotrimeric G-protein signaling pathway-Gq alpha and Go alpha mediated [50] | All | Age, Sex, 10PC | 1.00 (0.96, 1.04) | 0.872 |
| | Males | Age, 10PC | 0.97 (0.91, 1.02) | 0.214 |
| | <60 years | Sex, 10PC | 1.00 (0.95, 1.06) | 0.907 |
| | <60 years Males | 10PC | 0.99 (0.92, 1.06) | 0.737 |
| Heterotrimeric G-protein signaling pathway-Gi alpha and Gs alpha mediated [60] | All | Age, Sex, 10PC | 1.01 (0.97, 1.05) | 0.642 |
| | Males | Age, 10PC | 0.99 (0.93, 1.04) | 0.352 |
| | <60 years | Sex, 10PC | 1.03 (0.97, 1.09) | 0.320 |
| | <60 years Males | 10PC | 0.99 (0.92, 1.07) | 0.844 |

*SNPs reaching a P-value Threshold of $2.1 \times 10^{-3}$ (i.e., the best fit P-value Threshold when considering all hospitalised individuals). PC: Principal Components; SNP: single nucleotide polymorphism.

#### Supplementary Figures

**Figure S1. High-resolution PRSice-2 plot for the association of IPF PRS with COVID-19 hospitalisation.** The best-fit PRS is obtained at P-value Threshold (PT) of 2.10x10$^{-3}$, with a whole-genome PRS model including 2,939 variants from the GWAS of IPF dataset.

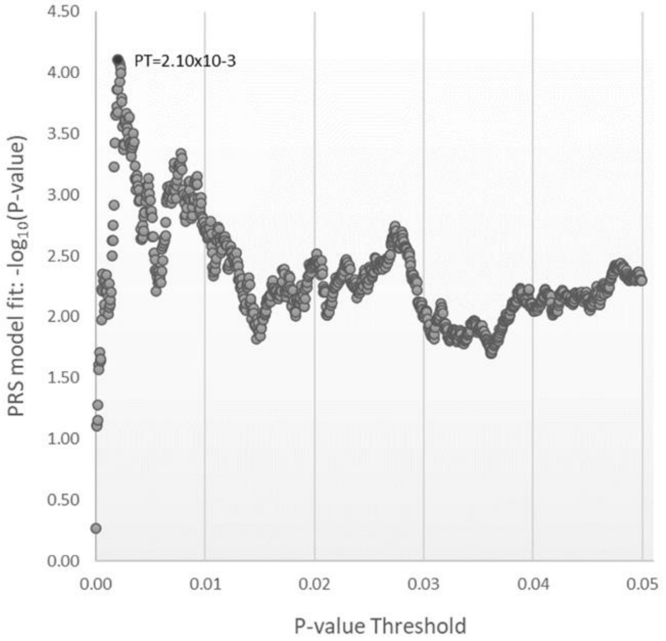

**Figure S2. Pathway enrichment analysis for genetic factors included in the best-fit model of association of IPF whole-genome PRS with COVID-19 hospitalisation for all individuals.** A) Analyses including all 2,939 SNPs used to calculate the PRS and their 3,471 mapping genes. B) Analyses excluding the 19 previously reported IPF common loci (sentinels ±1MB) (2,709 SNPs and 3,291 mapping genes left). All analyses were performed with ShinyGO v0.77 [5]. Top five biological pathways according to PANTHER database are shown.

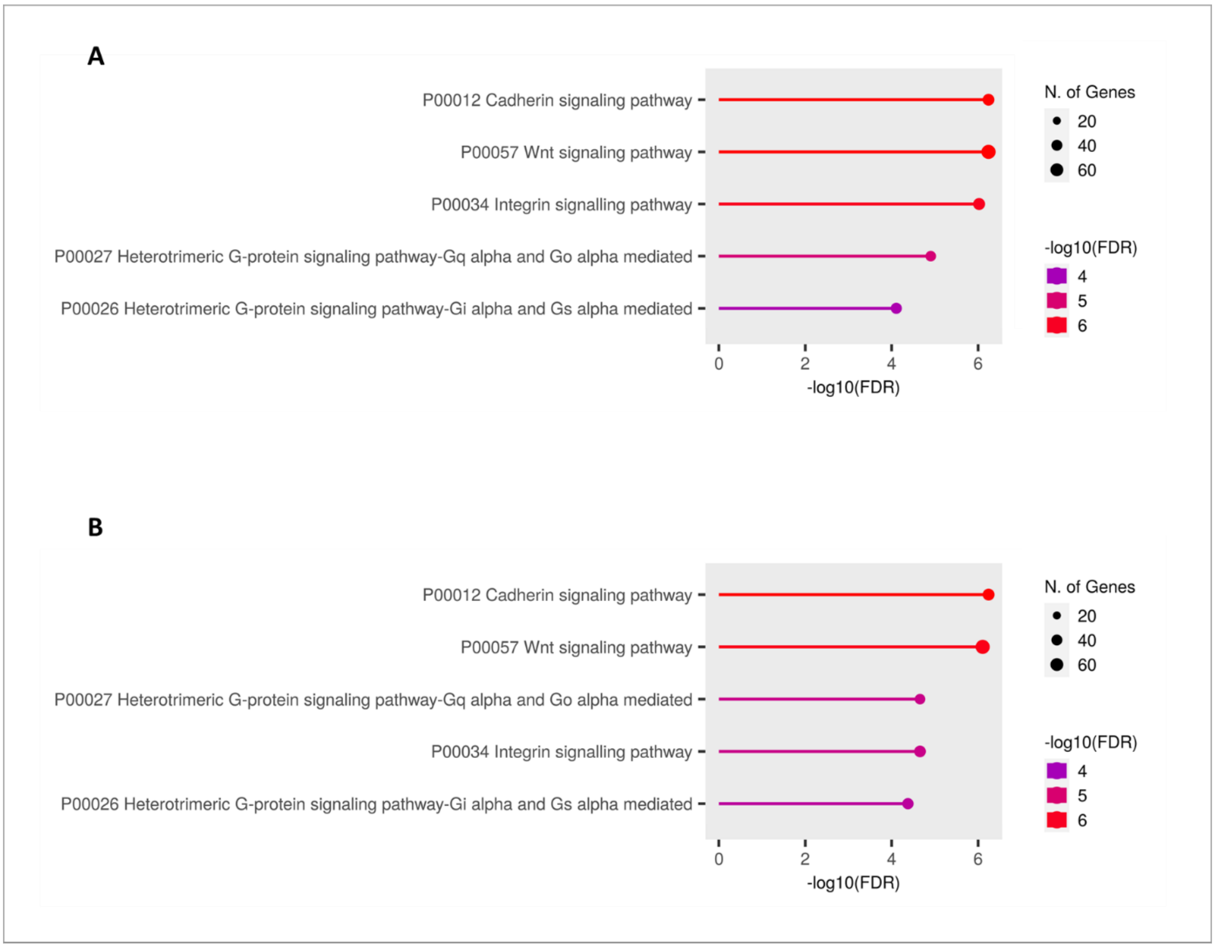

**Figure S3. Pathway enrichment analysis for genetic factors included in the best-fit model of association of IPF whole-genome PRS with COVID-19 hospitalisation for <60 years males.** Analyses including 5,660 SNPs used to calculate the PRS and their 5,424 mapping genes. Analyses were performed with ShinyGO v0.77 [5]. Top five biological pathways according to PANTHER database are shown.

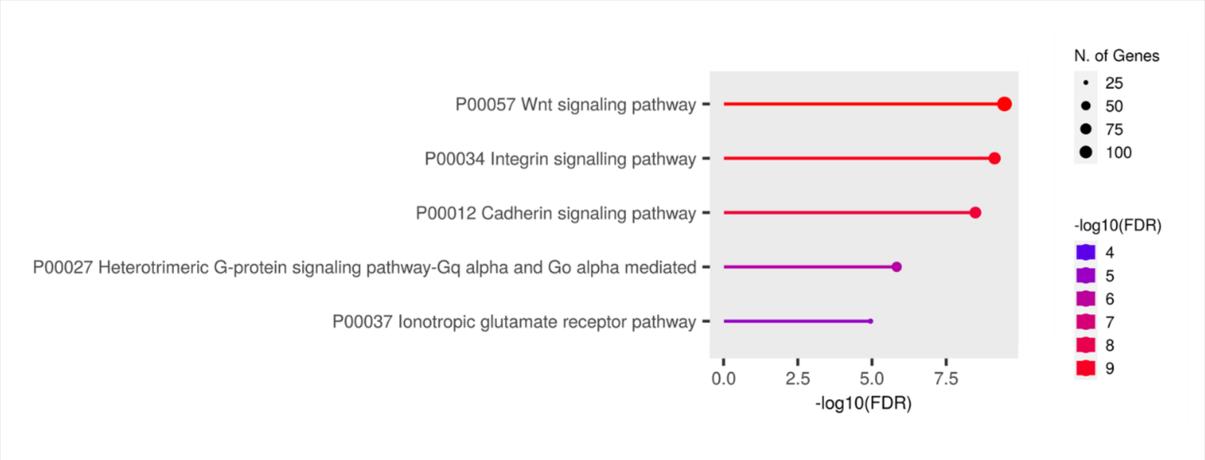

### Banner Scourge

26/09/2022

#### SCOURGE Cohort Group Members

Javier Abellan[1,2]; René Acosta-Isaac[3]; Jose María Aguado[4,5,6,7]; Carlos Aguilar[8]; Sergio Aguilera-Albesa[9,10]; Abdolah Ahmadi Sabbagh[11]; Jorge Alba[12]; Sergiu Albu[13,14,15]; Karla A.M. Alcalá-Gallardo[16]; Julia Alcoba-Florez[17]; Sergio Alcolea Batres[18]; Holmes Rafael Algarin-Lara[19,20]; Virginia Almadana[21]; Julia Almeida[22,23,24,25]; Berta Almoguera[26,27]; María R. Alonso[28]; Nuria Alvarez[28]; Yady Álvarez-Benítez[19,20]; Felipe Álvarez-Navia[29,30]; Rodolfo Alvarez-Sala Walther[18]; Álvaro Andreu-Bernabeu[31,6]; Maria Rosa Antonijoan[32]; Eunate Arana-Arri[33,34]; Carlos Aranda[35,36]; Celso Arango[31,37,6]; Carolina Araque[38,39]; Nathalia K. Araujo[40]; Izabel M.T. Araujo[41]; Ana C. Arcanjo[42,43,44]; Ana Arnaiz[45,46,47]; Francisco Arnalich Fernández[48]; María J. Arranz[49]; José Ramon Arribas Lopez[48]; Maria-Jesus Artiga[50]; Yubelly Avello-Malaver[51]; Carmen Ayuso[26,27]; Ana María Baldion[51]; Belén Ballina Martín[11]; Raúl C. Baptista-Rosas[52,53,54]; Andrea Barranco-Díaz[20]; María Barreda- Sánchez[55,56]; Viviana Barrera-Penagos[51]; Moncef Belhassen-Garcia[57,30]; Enrique Bernal[55]; David Bernal-Bello[58]; Joao F. Bezerra[59]; Marcos A.C. Bezerra[60]; Natalia Blanca-López[61]; Rafael Blancas[62]; Lucía Boix-Palop[63]; Alberto Borobia[64]; Elsa Bravo[65]; María Brion[66,67]; Óscar Brochado-Kith[68,7]; Ramón Brugada[69,70,67,71]; Matilde Bustos[72]; Alfonso Cabello[73]; Juan J. Caceres-Agra[74]; Esther Calbo[75]; Enrique J. Calderón[76,77,78]; Shirley Camacho[79]; Marcela C. Campos[42]; Yolanda Cañadas[36]; Cristina Carbonell[29,30]; Servando Cardona-Huerta[80]; Antonio Augusto F. Carioca[81]; Maria Sanchez Carpintero[35,36]; Carlos Carpio Segura[18]; Thássia M.T. Carratto[82]; José Antonio Carrillo-Avila[83]; Maria C.C. Carvalho[84]; Carlos Casasnovas[85,86,27]; Luis Castano[33,87,27,88,89]; Carlos F. Castaño[35,36]; Jose E. Castelao[90]; Aranzazu Castellano Candalija[91]; María A. Castillo[79]; Francisco C. Ceballos[68]; Jessica G. Chaux[39]; Walter G. Chaves- Santiago[92,39]; Sylena Chiquillo-Gómez[19,20]; Marco A. Cid-Lopez[16]; Oscar Cienfuegos-Jimenez[80]; Rosa Conde-Vicente[93]; M. Lourdes Cordero-Lorenzana[94]; Dolores Corella[95,96]; Almudena Corrales[97,98]; Jose L. Cortes-Sanchez[80,99]; Marta Corton[26,27]; Tatiana X. Costa[100]; Raquel Cruz[101,27,102,103]; Marina S. Cruz[40]; Luisa Cuesta[104]; Gabriela C.R. Cunha[105]; Gabriela V. da Silva[41]; David Dalmau[106,75]; Raquel C.S. Dantas-Komatsu[40]; M. Teresa Darnaude[107]; Raimundo de Andrés[108]; Jéssica N.G. de Araújo[109]; Carmen de Juan[110]; Juan De la Cruz Troca[111,112,77]; Carmen de la Horra[78]; Ana B. de la Hoz[33]; Alba De Martino-Rodríguez[113,114]; Julianna Lys de Sousa Alves Neri[115]; Victor del Campo-Pérez[116]; Juan Delgado-Cuesta[117]; Covadonga M. Diaz-Caneja[31,37,6]; Anderson Díaz-Pérez[20]; Aranzazu Diaz de Bustamante[107]; Beatriz Dietl[75]; Silvia Diz-de Almeida[27,103]; Manoella do Monte Alves[118,119]; Elena Domínguez-Garrido[120]; Katiusse A. dos Santos[84]; Alice M. Duarte[41]; Jose Echave-Sustaeta[121]; Rocío Eiros[122]; César O. Enciso-Olivera[38,39]; Gabriela Escudero[123]; Pedro Pablo España[124]; Gladys Mercedes Estigarribia Sanabria[125]; María Carmen Fariñas[45,46,47]; Marianne R. Fernandes[126,127]; Ramón Fernández[45,128]; Lidia Fernandez-Caballero[26,27]; Ana Fernández-Cruz[129]; María J. Fernandez-Nestosa[130]; Uxía Fernández-Robelo[131]; Amanda Fernández-Rodríguez[68,7]; Marta Fernández-Sampedro[45,47,46]; Ruth Fernández-Sánchez[26,27]; Tania Fernández-Villa[132]; Silvia Fernández Ferrero[11]; Yolanda Fernández Martínez[11]; Carmen Fernéndez Capitán[91]; Patricia Flores-Pérez[133]; Vicente Friaza[77,78]; Lácides Fuenmayor-Hernández[20]; Marta Fuertes Núñez[11]; Victoria Fumadó[134]; Ignacio Gadea[135]; Lidia Gagliardi[35,36]; Manuela Gago-Domínguez[136,102]; Natalia Gallego[137]; Cristina Galoppo[138]; Inés García[26,27]; Mercedes García[35,36]; Leticia García[35,36]; Carlos Garcia-Cerrada[1,2,139]; Aitor García-de-Vicuña[33,140]; Josefina Garcia-García[55]; Irene García-García[64]; Carmen García-Ibarbia[45,47,46]; Andrés C. García-Montero[141]; Ana García-Soidán[142]; Elisa García-Vázquez[55]; María Carmen García Torrejón[143,2]; Emiliano Garza-Frias[80]; Angela Gentile[138]; Belén Gil-Fournier[144]; Javier Gómez-Arrue[113,114]; Mario Gómez-Duque[92,39]; Luis Gómez Carrera[18]; María Gómez García[101]; Ángela Gómez Sacristán[145]; Anna González-Neira[28]; Javier González-Peñas[31,6,37]; Manuel Gonzalez-Sagrado[93]; Beatriz González Álvarez[113,114]; Fernan Gonzalez Bernaldo de Quirós[146]; Hugo Gonzalo Benito[147]; Oscar Gorgojo-Galindo[148]; Miguel Górgolas[73]; Florencia Guaragna[138]; Genilson P. Guegel[149]; Beatriz Guillen-Guio[97]; Encarna Guillen-Navarro[55,150,151,152]; Pablo Guisado-Vasco[121]; Juan F. Gutiérrez-Bautista[153]; Luz D. Gutierrez-Castañeda[154,39]; Sarah Heili-Frades[155]; Estefania Hernandez[156]; Luis D. Hernandez-Ortega[157,158]; Guillermo Hernández-Pérez[29]; Rebeca Hernández-Vaquero[159]; Cristina Hernández Moro[11]; Belen

Herraez[28]; M. Teresa Herranz[55]; María Herrera[35,36]; María José Herrero[160,161]; Antonio Herrero-Gonzalez[162]; Juan P. Horcajada[163,164,14,165,7]; Natale Imaz-Ayo[33]; Maider Intxausti-Urrutibeaskoa[166]; María Íñiguez[167]; Rafael H. Jacomo[168]; Rubén Jara[55]; Perez Maria Jazmin[138]; Ángel Jiménez[35,36]; Pilar Jiménez[153]; Ignacio Jiménez-Alfaro[169]; María A. Jimenez-Sousa[68,7]; Iolanda Jordan[170,171,77]; Rocío Laguna-Goya[172,173]; Daniel Laorden[18]; María Lasa-Lazaro[172,173]; María Claudia Lattig[79,174]; Ailen Lauriente[138]; Anabel Liger Borja[175]; Lucía Llanos[176]; Amparo López-Bernús[29,30]; Esther Lopez-Garcia[111,112,77,177]; Rosario Lopez-Rodriguez[26,27]; Miguel A. López-Ruz[178,179,180]; Eduardo López Granados[181,182,27]; Leonardo Lorente[183]; José E. Lozano[184]; María Lozano-Espinosa[175]; Andre D. Luchessi[185]; Ignacio Mahillo[186,187,98]; Esther Mancebo[172,173]; Carmen Mar[124]; Cristina Marcelo Calvo[91]; Miguel Marcos[29,30]; Alba Marcos-Delgado[188]; Alicia Marín Candon[64]; Pablo Mariscal Aguilar[18]; María M. Martín[189]; María Dolores Martín[190]; Vicente Martín[188,77]; Marta Martin-Fernandez[191]; Caridad Martín-López[175]; José-Ángel Martín-Oterino[29,30]; Laura Martin-Pedraza[61]; María Martín-Vicente[68]; Amalia Martinez[192]; Ricardo Martínez[156]; Juan José Martínez[86,27]; Silvia Martínez[45,47]; Eleno Martínez-Aquino[193]; Óscar Martínez-González[194]; Iciar Martinez-Lopez[195,196]; Oscar Martinez-Nieto[51,174]; Pedro Martinez-Paz[147]; Angel Martinez-Perez[197]; Andrea Martínez-Ramas[26,27]; Michel F. Martinez-Resendez[80]; Violeta Martínez Robles[11]; Laura Marzal[26,27]; Juliana F. Mazzeu[198,199,200]; Jeane F.P. Medeiros[40]; Kelliane A. Medeiros[201,202]; Francisco J. Medrano[76,77,78]; Xose M. Meijome[203,204]; Natalia Mejuto-Montero[205]; Ana Méndez-Echevarria[206]; Humberto Mendoza Charris[65,20]; Eleuterio Merayo Macías[207]; Fátima Mercadillo[208]; Arieh R. Mercado-Sesma[157,158]; Pablo Minguez[26,27]; Antonio J J. Molina[188,77]; Elena Molina-Roldán[209]; Juan José Montoya[156]; Vitor M.S. Moraes[82]; Patricia Moreira-Escriche[110]; Xenia Morelos-Arnedo[65,20]; Antonio Moreno-Docón[55]; Junior Moreno-Escalante[20]; Victor Moreno Cuerda[1,2]; Alberto Moreno Fernández[91]; Rubén Morilla[78,210]; Patricia Muñoz García[211,98,6]; Pablo Neira[138]; Julian Nevado[27,137,212]; Israel Nieto-Gañán[142]; Joana F.R. Nunes[42]; Rocio Nuñez- Torres[28]; Antònia Obrador-Hevia[213,214]; J. Gonzalo Ocejo-Vinyals[45,47]; Virginia Olivar[138]; Silviene F. Oliveira[198,215,216,217,218]; Lorena Ondo[26,27]; Alberto Orfao[22,23,24,25]; Luis Ortega[219]; Eva Ortega-Paino[50]; Fernando Ortiz-Flores[45,47]; Rocio Ortiz-Lopez[220,80]; José A. Oteo[12,167]; Harry Pachajoa[221,222]; Manuel Pacheco[156]; Fredy Javier Pacheco-Miranda[20]; Irene Padilla Conejo[11]; Sonia Panadero-Fajardo[83]; Mara Parellada[31,37,6]; Roberto Pariente-Rodríguez[142]; Estela Paz-Artal[172,173,223]; Germán Peces-Barba[224,98]; Miguel S. Pedromingo Kus[225]; Celia Perales[135]; Patricia Perez[226]; César Pérez[227]; Gustavo Perez-de-Nanclares[33,87]; Felipe Pérez-García[228,229]; Patricia Pérez-Matute[167]; Alexandra Pérez-Serra[69,67]; M. Elena Pérez-Tomás[55]; Teresa Perucho[230]; Lisbeth A. Pichardo[11]; Susana M.T. Pinho[201,231,232]; Mel·lina Pinsach-Abuin[69,67]; Luz Adriana Pinzón[92,39]; Guillermo Pita[28]; Francesc Pla-Junca[233,27]; Laura Planas-Serra[86,27]; Ericka N. Pompa-Mera[234]; Gloria L. Porras-Hurtado[156]; Aurora Pujol[86,27,235]; María Eugenia Quevedo Chávez[19,20]; Maria Angeles Quijada[32,236]; Inés Quintela[101]; Diana Ramirez-Montaño[237]; Soraya Ramiro León[144]; Pedro Rascado Sedes[238]; Delia Recalde[113,114]; Emma Recio-Fernández[167]; Salvador Resino[68,7]; Adriana P. Ribeiro[201,202,232]; Carlos S. Rivadeneira-Chamorro[39]; Diana Roa-Agudelo[51]; Montserrat Robelo Pardo[238]; Marilyn Johanna Rodriguez[39]; Fernando Rodriguez-Artalejo[111,112,77,177]; Marena Rodríguez-Ferrer[20]; Carlos Rodriguez-Gallego[239,240]; José A. Rodriguez-Garcia[11]; María A. Rodriguez-Hernandez[72]; Antonio Rodriguez-Nicolas[153]; Agustí Rodriguez-Palmero[241,86]; Emilio Rodríguez-Ruiz[238,102]; Paula A. Rodriguez-Urrego[51]; Belén Rodríguez Maya[1]; German Ezequiel Rodriguez Novoa[138]; Federico Rojo[242,25]; Andrea Romero-Coronado[20]; Filomeno Rondón García[11]; Lidia S. Rosa[243]; Antonio Rosales-Castillo[244]; Cladelis Rubio[245,246]; María Rubio Olivera[35,36]; Montserrat Ruiz[86,27]; Francisco Ruiz-Cabello[153,179,247]; Eva Ruiz-Casares[230]; Juan J. Ruiz-Cubillan[45,47]; Javier Ruiz-Hornillos[248,36,249]; Pablo Ryan[250,251,252]; Hector D. Salamanca[38,39]; Lorena Salazar-García[79]; Giorgina Gabriela Salgueiro Origlia [91]; Pedro-Luis Sánchez[122,30]; Clara Sánchez-Pablo[122]; Olga Sánchez-Pernaute[253]; Antonio J. Sánchez López[254]; María Concepción Sánchez Prados[18]; Javier Sánchez Real[11]; Jorge Sánchez Redondo[1,255]; Cristina Sancho- Sainz[166]; Anna Sangil[63]; Arnoldo Santos[227]; Ney P.C. Santos[126]; Agatha Schlüter[86,27]; Sonia Segovia[233,256,257]; Alex Serra-Llovich[258]; Fernando Sevil Puras[8]; Marta Sevilla Porras[27,137]; Miguel A. Sicolo[259,260]; Vivian N. Silbiger[185]; Nayara S. Silva[109]; Fabiola T.C. Silva[42]; Cristina Silván Fuentes[27]; Jordi Solé-Violán[261,98,262]; José Manuel Soria[197]; Jose V. Sorlí[95,96]; Renata R. Sousa[198]; Juan Carlos Souto[3]; Karla S.C. Souza[84]; Vanessa S. Souza[105]; John J. Sprockel[92,39]; José Javier Suárez-Rama[101]; David A.

Suarez-Zamora[51]; Xiana Taboada-Fraga[205]; Eduardo Tamayo[263,148]; Alvaro Tamayo-Velasco[264]; Juan Carlos Taracido-Fernandez[162]; Nathali A.C. Tavares[265]; Carlos Tellería[113,114]; Jair Antonio Tenorio Castaño[27,137,212]; Alejandro Teper[138]; Juan Torres-Macho[266]; Lilian Torres-Tobar[39]; Ronald P. Torres Gutiérrez[225]; Jesús Troya[250]; Miguel Urioste[208]; Juan Valencia-Ramos[267]; Agustín Valido[21,268]; Juan Pablo Vargas Gallo[269,270]; Belén Varón[271]; Romero H.T. Vasconcelos[265]; Tomas Vega[272]; Santiago Velasco-Quirce[273]; Valentina Vélez-Santamaría[85,86]; Virginia Víctor[35,36]; Julia Vidán Estévez[11]; Miriam Vieitez-Santiago[45,47]; Carlos Vilches[274]; Lavinia Villalobos[11]; Felipe Villar[224]; Judit Villar-Garcia[275,276,277]; Cristina Villaverde[26,27]; Pablo Villoslada-Blanco[167]; Ana Virseda-Berdices[68]; Zuleima Yáñez[20]; Antonio Zapatero-Gaviria[278]; Ruth Zarate[279]; Sandra Zazo[242]; Miguel López de Heredia[27]; Ingrid Mendes[27]; Rocío Moreno[27]; Esther Sande[27,102,103]; Carlos Flores[280,97,98,240]; José A. Riancho[45,46,47]; Augusto Rojas-Martinez[80]; Pablo Lapunzina[27,137,212]; Angel Carracedo[101,27,102,103,136]

#### SCOURGE Cohort Group Affiliations

[1], Hospital Universitario Mostoles, Medicina Interna, Madrid, Spain
[2], Universidad Francisco de Vitoria, Madrid, Spain
[3], Haemostasis and Thrombosis Unit, Hospital de la Santa Creu i Sant Pau, IIB Sant Pau, Barcelona, Spain
[4], Unit of Infectious Diseases, Hospital Universitario 12 de Octubre, Instituto de Investigación Sanitaria Hospital 12 de Octubre (imas12), Madrid, Spain
[5], Spanish Network for Research in Infectious Diseases (REIPI RD16/0016/0002), Instituto de Salud Carlos III, Madrid, Spain
[6], School of Medicine, Universidad Complutense, Madrid, Spain
[7], Centro de Investigación Biomédica en Red de Enfermedades Infecciosas (CIBERINFEC), Instituto de Salud Carlos III, Madrid, Spain
[8], Hospital General Santa Bárbara de Soria, Soria, Spain
[9], Pediatric Neurology Unit, Department of Pediatrics, Navarra Health Service Hospital, Pamplona, Spain
[10], Navarra Health Service, NavarraBioMed Research Group, Pamplona, Spain
[11], Complejo Asistencial Universitario de León, León, Spain
[12], Hospital Universitario San Pedro, Infectious Diseases Department, Logroño, Spain
[13], Fundación Institut Guttmann, Institut Universitari de Neurorehabilitació adscrit a la UAB, Hospital de Neurorehabilitació, Barcelona, Spain
[14], Universitat Autònoma de Barcelona (UAB), Barcelona, Spain
[15], Fundació Institut d'Investigació en Ciències de la Salut Germans Trias i Pujol, Barcelona, Spain
[16], Hospital General de Occidente, Guadalajara, Mexico
[17], Microbiology Unit, Hospital Universitario N. S. de Candelaria, Santa Cruz de Tenerife, Spain
[18], Hospital Universitario La Paz-IDIPAZ, Servicio de Neumología, Madrid, Spain
[19], Camino Universitario Adelita de Char, Mired IPS, Barranquilla, Colombia
[20], Universidad Simón Bolívar, Facultad de Ciencias de la Salud, Barranquilla, Colombia
[21], Hospital Universitario Virgen Macarena, Neumología, Seville, Spain
[22], Departamento de Medicina, Universidad de Salamanca, Salamanca, Spain
[23], Centro de Investigación del Cáncer (IBMCC) Universidad de Salamanca - CSIC, Salamanca, Spain
[24], Biomedical Research Institute of Salamanca (IBSAL) Salamanca, Spain
[25], Centre for Biomedical Network Research on Cancer (CIBERONC), Instituto de Salud Carlos III, Madrid, Spain
[26], Department of Genetics & Genomics, Instituto de Investigación Sanitaria-Fundación Jiménez Díaz University Hospital - Universidad Autónoma de Madrid (IIS-FJD, UAM), Madrid, Spain
[27], Centre for Biomedical Network Research on Rare Diseases (CIBERER), Instituto de Salud Carlos III, Madrid, Spain
[28], Spanish National Cancer Research Centre, Human Genotyping-CEGEN Unit, Madrid, Spain

[29], Hospital Universitario de Salamanca-IBSAL, Servicio de Medicina Interna, Salamanca, Spain
[30], Universidad de Salamanca, Salamanca, Spain
[31], Department of Child and Adolescent Psychiatry, Institute of Psychiatry and Mental Health, Hospital General Universitario Gregorio Marañón (IiSGM), Madrid, Spain
[32], Clinical Pharmacology Service, Hospital de la Santa Creu i Sant Pau, IIB Sant Pau, Barcelona, Spain
[33], Biocruces Bizkai HRI, Barakaldo, Bizkaia, Spain
[34], Cruces University Hospital, Osakidetza, Barakaldo, Bizkaia, Spain
[35], Hospital Infanta Elena, Valdemoro, Madrid, Spain
[36], Instituto de Investigación Sanitaria-Fundación Jiménez Díaz University Hospital - Universidad Autónoma de Madrid (IIS-FJD, UAM), Madrid, Spain
[37], Centre for Biomedical Network Research on Mental Health (CIBERSAM), Instituto de Salud Carlos III, Madrid, Spain
[38], Fundación Hospital Infantil Universitario de San José, Bogotá, Colombia
[39], Fundación Universitaria de Ciencias de la Salud, Bogotá, Colombia
[40], Universidade Federal do Rio Grande do Norte, Programa de Pós-graduação em Ciências da Saúde, Natal, Brazil
[41], Universidade Federal do Rio Grande do Norte, Departamento de Medicina Clínica, Natal, Brazil
[42], Departamento de Genética e Morfologia, Instituto de Ciências Biológicas, Universidade de Brasília, Brasilia, Brazil
[43], Colégio Marista de Brasilia, Brazil
[44], Associação Brasileira de Educação e Cultura, Brazil
[45], IDIVAL, Santander, Spain
[46], Universidad de Cantabria, Santander, Spain
[47], Hospital U M Valdecilla, Santander, Spain
[48], Hospital Universitario La Paz-IDIPAZ, Servicio de Medicina Interna, Madrid, Spain
[49], Fundació Docència I Recerca Mutua Terrassa, Barcelona, Spain
[50], Spanish National Cancer Research Center, CNIO Biobank, Madrid, Spain
[51], Fundación Santa Fe de Bogota, Departamento Patologia y Laboratorios, Bogotá, Colombia
[52], Hospital General de Occidente, Zapopan, Jalisco, Mexico
[53], Centro Universitario de Tonalá, Universidad de Guadalajara, Tonalá, Jalisco, Mexico
[54], Centro de Investigación Multidisciplinario en Salud, Universidad de Guadalajara, Tonalá, Jalisco, Mexico
[55], Instituto Murciano de Investigación Biosanitaria (IMIB-Arrixaca), Murcia, Spain
[56], Universidad Católica San Antonio de Murcia (UCAM), Murcia, Spain
[57], Hospital Universitario de Salamanca-IBSAL, Servicio de Medicina Interna-Unidad de Enfermedades Infecciosas, Salamanca, Spain
[58], Hospital Universitario de Fuenlabrada, Department of Internal Medicine, Madrid, Spain
[59], Escola Tecnica de Saúde, Laboratorio de Vigilancia Molecular Aplicada, Pará, Brazil
[60], Federal University of Pernambuco, Genetics Postgraduate Program, Recife, PE, Brazil
[61], Hospital Universitario Infanta Leonor, Servicio de Alergia, Madrid, Spain
[62], Hospital Universitario del Tajo, Servicio de Medicina Intensiva, Aranjuez, Spain
[63], Hospital Universitario Mutua Terrassa, Barcelona, Spain
[64], Hospital Universitario La Paz-IDIPAZ, Servicio de Farmacología, Madrid, Spain
[65], Alcaldía de Barranquilla, Secretaría de Salud, Barranquilla, Colombia
[66], Instituto de Investigación Sanitaria de Santiago (IDIS), Xenética Cardiovascular, Santiago de Compostela, Spain
[67], Centre for Biomedical Network Research on Cardiovascular Diseases (CIBERCV), Instituto de Salud Carlos III, Madrid, Spain
[68], Unidad de Infección Viral e Inmunidad, Centro Nacional de Microbiología (CNM), Instituto de Salud Carlos III (ISCIII), Madrid, Spain
[69], Cardiovascular Genetics Center, Institut d’Investigació Biomèdica Girona (IDIBGI), Girona, Spain

[70], Medical Science Department, School of Medicine, University of Girona, Girona, Spain
[71], Hospital Josep Trueta, Cardiology Service, Girona, Spain
[72], Institute of Biomedicine of Seville (IBiS), Consejo Superior de Investigaciones Científicas (CSIC)-University of Seville- Virgen del Rocio University Hospital, Seville, Spain
[73], Division of Infectious Diseases, Instituto de Investigación Sanitaria-Fundación Jiménez Díaz University Hospital - Universidad Autónoma de Madrid (IIS-FJD, UAM), Madrid, Spain
[74], Intensive Care Unit, Hospital Universitario Insular de Gran Canaria, Las Palmas de Gran Canaria, Spain
[75], Hospital Universitario Mutua Terrassa, Terrassa, Spain
[76], Departemento de Medicina, Hospital Universitario Virgen del Rocío,Universidad de Sevilla, Seville, Spain
[77], Centre for Biomedical Network Research on Epidemiology and Public Health (CIBERESP), Instituto de Salud Carlos III, Madrid, Spain
[78], Instituto de Biomedicina de Sevilla, Seville, Spain
[79], Universidad de los Andes, Facultad de Ciencias, Bogotá, Colombia
[80], Tecnologico de Monterrey, Escuela de Medicina y Ciencias de la Salud and Hospital San Jose TecSalud, Monterrey, Mexico
[81], University of Fortaleza (UNIFOR), Department of Nutrition. Fortaleza, Brazil
[82], Departamento de Química, Faculdade de Filosofia, Ciências e Letras de Ribeirão Preto, Universidade de São Paulo, Brazil
[83], Andalusian Public Health System Biobank, Granada, Spain
[84], Universidade Federal do Rio Grande do Norte, Programa de Pós-Graduação em Ciências Farmacêuticas, Natal, Brazil
[85], Neuromuscular Unit, Neurology Department, Hospital Universitari de Bellvitge, L'Hospitalet de Llobregat (Barcelona), Spain
[86], Bellvitge Biomedical Research Institute (IDIBELL), Neurometabolic Diseases Laboratory, L'Hospitalet de Llobregat, Spain
[87], Osakidetza, Cruces University Hospital, Barakaldo, Bizkaia, Spain
[88], Centre for Biomedical Network Research on Diabetes and Metabolic Associated Diseases (CIBERDEM), Instituto de Salud Carlos III, Madrid, Spain
[89], University of Pais Vasco, UPV/EHU, Bizkaia, Spain
[90], Oncology and Genetics Unit, Instituto de Investigacion Sanitaria Galicia Sur, Xerencia de Xestion Integrada de Vigo-Servizo Galego de Saúde, Vigo, Spain
[91], Hospital Universitario La Paz, Hospital Carlos III, Madrid, Spain
[92], Hospital de San José, Sociedad de Cirugía de Bogota, Bogotá, Colombia
[93], Hospital Universitario Río Hortega, Valladolid, Spain
[94], Servicio de Medicina intensiva, Complejo Hospitalario Universitario de A Coruña (CHUAC), Sistema Galego de Saúde (SERGAS), A Coruña, Spain
[95], Valencia University, Preventive Medicine Department, Valencia, Spain
[96], Centre for Biomedical Network Research on Physiopatology of Obesity and Nutrition (CIBEROBN), Instituto de Salud Carlos III, Madrid, Spain
[97], Research Unit, Hospital Universitario N.S. de Candelaria, Santa Cruz de Tenerife, Spain
[98], Centre for Biomedical Network Research on Respiratory Diseases (CIBERES), Instituto de Salud Carlos III, Madrid, Spain
[99], Otto von Guericke University, Departament of Microgravity and Translational Regenerative Medicine, Magdeburg, Germany
[100], Maternidade Escola Janário Cicco, Natal, Brazil
[101], Centro Nacional de Genotipado (CEGEN), Universidade de Santiago de Compostela, Santiago de Compostela, Spain
[102], Instituto de Investigación Sanitaria de Santiago (IDIS), Santiago de Compostela, Spain

[103], Centro Singular de Investigación en Medicina Molecular y Enfermedades Crónicas (CIMUS), Universidade de Santiago de Compostela, Santiago de Compostela, Spain
[104], Institute of Psychiatry and Mental Health, Hospital General Universitario Gregorio Marañón (IiSGM), Madrid, Spain
[105], Programa de Pós Graduação em Ciências da Saúde, Faculdade de Medicina, Universidade de Brasília, Brasilia, Brazil
[106], Fundació Docència I Recerca Mutua Terrassa, Terrassa, Spain
[107], Hospital Universitario Mostoles, Unidad de Genética, Madrid, Spain
[108], Internal Medicine Department, Instituto de Investigación Sanitaria-Fundación Jiménez Díaz University Hospital - Universidad Autónoma de Madrid (IIS-FJD, UAM), Madrid, Spain
[109], Universidade Federal do Rio Grande do Norte, Pós-graduação em Biotecnologia - Rede de Biotecnologia do Nordeste (Renorbio), Natal, Brazil
[110], Hospital Universitario Severo Ochoa, Servicio de Medicina Interna, Madrid, Spain
[111], Department of Preventive Medicine and Public Health, School of Medicine, Universidad Autónoma de Madrid, Madrid, Spain
[112], IdiPaz (Instituto de Investigación Sanitaria Hospital Universitario La Paz), Madrid, Spain
[113], Instituto Aragonés de Ciencias de la Salud (IACS), Zaragoza, Spain
[114], Instituto Investigación Sanitaria Aragón (IIS-Aragon), Zaragoza, Spain
[115], Universidade Federal do Rio Grande do Norte, Programa de Pós Graduação em Nutrição, Natal, Brazil
[116], Preventive Medicine Department, Instituto de Investigacion Sanitaria Galicia Sur, Xerencia de Xestion Integrada de Vigo-Servizo Galego de Saúde, Vigo, Spain
[117], Hospital Universitario Virgen del Rocío, Servicio de Medicina Interna, Seville, Spain
[118], Universidade Federal do Rio Grande do Norte, Departamento de Infectologia, Natal, Brazil
[119], Hospital de Doenças Infecciosas Giselda Trigueiro, Rio Grande do Norte, Natal, Brazil
[120], Unidad Diagnóstico Molecular. Fundación Rioja Salud, La Rioja, Spain
[121], Hospital Universitario Quironsalud Madrid, Madrid, Spain
[122], Hospital Universitario de Salamanca-IBSAL, Servicio de Cardiología, Salamanca, Spain
[123], Hospital Universitario Puerta de Hierro, Servicio de Medicina Interna, Majadahonda, Spain
[124], Biocruces Bizkaia Health Research Institute, Galdakao University Hospital, Osakidetza, Bizkaia, Spain
[125], Instituto Regional de Investigación en Salud-Universidad Nacional de Caaguazú, Caaguazú, Paraguay
[126], Universidade Federal do Pará, Núcleo de Pesquisas em Oncologia, Belém, Pará, Brazil
[127], Hospital Ophir Loyola, Departamento de Ensino e Pesquisa, Belém, Pará, Brazil
[128], Fundación Asilo San Jose, Santander, Spain
[129], Unidad de Enfermedades Infecciosas, Servicio de Medicina Interna, Hospital Universitario Puerta de Hierro, Instituto de Investigación Sanitaria Puerta de Hierro - Segovia de Arana, Madrid, Spain
[130], Universidad Nacional de Asunción, Facultad de Politécnica, Paraguay
[131], Urgencias Hospitalarias, Complejo Hospitalario Universitario de A Coruña (CHUAC), Sistema Galego de Saúde (SERGAS), A Coruña, Spain
[132], Grupo de Investigación en Interacciones Gen-Ambiente y Salud (GIIGAS) - Instituto de Biomedicina (IBIOMED), Universidad de León, León, Spain
[133], Hospital Universitario Niño Jesús, Pediatrics Department, Madrid, Spain
[134], Unitat de Malalties Infeccioses i Importades, Servei de Pediatría, Infectious and Imported Diseases, Pediatric Unit, Hospital Universitari Sant Joan de Deú, Barcelona, Spain
[135], Microbiology Department, Instituto de Investigación Sanitaria-Fundación Jiménez Díaz University Hospital - Universidad Autónoma de Madrid (IIS-FJD, UAM), Madrid, Spain
[136], Fundación Pública Galega de Medicina Xenómica, Sistema Galego de Saúde (SERGAS) Santiago de Compostela, Spain

[137], Instituto de Genética Médica y Molecular (INGEMM), Hospital Universitario La Paz-IDIPAZ, Madrid, Spain
[138], Hospital de Niños Ricardo Gutierrez, Buenos Aires, Argentina
[139], Centre for Biomedical Network Research on Rare Diseases (CIBERER), Instituto de Salud Carlos III, Madrid, Spain Universidad Francisco de Vitoria, Madrid,Spain
[140], Osakidetza, Cruces University Hospital, Bizkaia, Spain
[141], University of Salamanca, Biomedical Research Institute of Salamanca (IBSAL), Salamanca, Spain
[142], Department of Immunology, IRYCIS, Hospital Universitario Ramón y Cajal, Madrid, Spain
[143], Hospital Infanta Elena, Servicio de Medicina Intensiva, Valdemoro, Madrid, Spain
[144], Hospital Universitario de Getafe, Servicio de Genética, Madrid, Spain
[145], Pneumology Department, Hospital General Universitario Gregorio Marañón (iiSGM), Madrid, Spain
[146], Ministerio de Salud Ciudad de Buenos Aires, Buenos Aires, Argentina
[147], Hospital Clinico Universitario de Valladolid, Unidad de Apoyo a la Investigación, Valladolid, Spain
[148], Universidad de Valladolid, Departamento de Cirugía, Valladolid, Spain
[149], Secretaria Municipal de Saude de Apodi, Natal, Brazil
[150], Sección Genética Médica - Servicio de Pediatría, Hospital Clínico Universitario Virgen de la Arrixaca, Servicio Murciano de Salud, Murcia, Spain
[151], Departamento Cirugía, Pediatría, Obstetricia y Ginecología, Facultad de Medicina, Universidad de Murcia (UMU), Murcia, Spain
[152], Grupo Clínico Vinculado, Centre for Biomedical Network Research on Rare Diseases (CIBERER), Instituto de Salud Carlos III, Madrid, Spain
[153], Hospital Universitario Virgen de las Nieves, Servicio de Análisis Clínicos e Inmunología, Granada, Spain
[154], Hospital Universitario Centro Dermatológico Federico Lleras Acosta, Bogotá, Colombia
[155], Intermediate Respiratory Care Unit, Department of Pneumology, Instituto de Investigación Sanitaria-Fundación Jiménez Díaz University Hospital - Universidad Autónoma de Madrid (IIS-FJD, UAM), Madrid, Spain
[156], Clinica Comfamiliar Risaralda, Pereira, Colombia
[157], Centro Universitario de Tonalá, Universidad de Guadalajara, Guadalajara, Mexico
[158], Centro de Investigación Multidisciplinario en Salud, Universidad de Guadalajara, Guadalajara, Mexico
[159], Unidad de Cuidados, Intensivos Hospital Clínico Universitario de Santiago (CHUS), Sistema Galego de Saúde (SERGAS), Santiago de Compostela, Spain
[160], IIS La Fe, Plataforma de Farmacogenética, Valencia, Spain
[161], Universidad de Valencia, Departamento de Farmacología, Valencia, Spain
[162], Data Analysis Department, Instituto de Investigación Sanitaria-Fundación Jiménez Díaz University Hospital - Universidad Autónoma de Madrid (IIS-FJD, UAM), Madrid, Spain
[163], Hospital del Mar, Infectious Diseases Service, Barcelona, Spain
[164], Institut Hospital del Mar d'Investigacions Mèdiques (IMIM), Barcelona, Spain
[165], CEXS-Universitat Pompeu Fabra, Spanish Network for Research in Infectious Diseases (REIPI), Barcelona, Spain
[166], Biocruces Bizkaia Health Research Institute, Basurto University Hospital, Osakidetza, Bizkaia, Spain
[167], Infectious Diseases, Microbiota and Metabolism Unit, Center for Biomedical Research of La Rioja (CIBIR), Logroño, Spain
[168], Sabin Medicina Diagnóstica, Brazil
[169], Opthalmology Department, Instituto de Investigación Sanitaria-Fundación Jiménez Díaz University Hospital - Universidad Autónoma de Madrid (IIS-FJD, UAM), Madrid, Spain
[170], Hospital Sant Joan de Deu,Pediatric Critical Care Unit, Barcelona, Spain
[171], Paediatric Intensive Care Unit, Agrupación Hospitalaria Clínic-Sant Joan de Déu, Esplugues de Llobregat, Barcelona, Spain
[172], Hospital Universitario 12 de Octubre, Department of Immunology, Madrid, Spain

[173], Instituto de Investigación Sanitaria Hospital 12 de Octubre (imas12), Transplant Immunology and Immunodeficiencies Group, Madrid, Spain
[174], SIGEN Alianza Universidad de los Andes - Fundación Santa Fe de Bogotá, Bogotá, Colombia
[175], Hospital General de Segovia, Medicina Intensiva, Segovia, Spain
[176], Clinical Trials Unit, Instituto de Investigación Sanitaria-Fundación Jiménez Díaz University Hospital - Universidad Autónoma de Madrid (IIS-FJD, UAM), Madrid, Spain
[177], IMDEA-Food Institute, CEI UAM+CSIC, Madrid, Spain
[178], Hospital Universitario Virgen de las Nieves, Servicio de Enfermedades Infecciosas, Granada, Spain
[179], Instituto de Investigación Biosanitaria de Granada (ibs GRANADA), Granada, Spain
[180], Universidad de Granada, Departamento de Medicina, Granada, Spain
[181], Hospital Universitario La Paz-IDIPAZ, Servicio de Inmunología, Madrid, Spain
[182], La Paz Institute for Health Research (IdiPAZ), Lymphocyte Pathophysiology in Immunodeficiencies Group, Madrid, Spain
[183], Intensive Care Unit, Hospital Universitario de Canarias, La Laguna, Spain
[184], Dirección General de Salud Pública, Consejería de Sanidad, Junta de Castilla y León, Valladolid, Spain
[185], Universidade Federal do Rio Grande do Norte, Departamento de Analises Clinicas e Toxicologicas, Natal, Brazil
[186], Fundación Jiménez Díaz, Epidemiology, Madrid, Spain
[187], Universidad Autónoma de Madrid, Department of Medicine, Madrid, Spain
[188], Instituto de Biomedicina (IBIOMED), Universidad de León, León, Spain
[189], Intensive Care Unit, Hospital Universitario N. S. de Candelaria, Santa Cruz de Tenerife, Spain
[190], Preventive Medicine Department, Instituto de Investigación Sanitaria-Fundación Jiménez Díaz University Hospital - Universidad Autónoma de Madrid (IIS-FJD, UAM), Madrid, Spain
[191], Universidad de Valladolid, Departamento de Medicina, Valladolid, Spain
[192], Hospital Universitario Infanta Leonor, Servicio de Medicina Intensiva, Madrid, Spain
[193], Servicio de Medicina Interna, Sanatorio Franchin, Buenos Aires, Argentina
[194], Hospital Universitario del Tajo, Servicio de Medicina Intensiva, Toledo, Spain
[195], Unidad de Genética y Genómica Islas Baleares, Islas Baleares, Spain
[196], Hospital Universitario Son Espases, Unidad de Diagnóstico Molecular y Genética Clínica, Islas Baleares, Spain
[197], Genomics of Complex Diseases Unit, Research Institute of Hospital de la Santa Creu i Sant Pau, IIB Sant Pau, Barcelona, Spain
[198], Faculdade de Medicina, Universidade de Brasília, Brasilia, Brazil
[199], Programa de Pós-Graduação em Ciências Médicas, Universidade de Brasília, Brasilia, Brazil
[200], Programa de Pós-Graduação em Ciências da Saúde, Universidade de Brasília, Brasilia, Brazil
[201], Hospital das Forças Armadas, Brazil
[202], Exército Brasileiro, Brazil
[203], Hospital El Bierzo, Gerencia de Asistencia Sanitaria del Bierzo (GASBI), Gerencia Regional de Salud (SACYL), Ponferrada, Spain
[204], Grupo INVESTEN, Instituto de Salud Carlos III, Madrid, Spain
[205], Unidad de Cuidados Intensivos, Complejo Universitario de A Coruña (CHUAC), Sistema Galego de Saúde (SERGAS), A Coruña, Spain
[206], Hospital Universitario La Paz-IDIPAZ, Servicio de Pediatría, Madrid, Spain
[207], Hospital El Bierzo, Unidad Cuidados Intensivos, León, Spain
[208], Spanish National Cancer Research Centre, Familial Cancer Clinical Unit, Madrid, Spain
[209], Instituto de Investigación Sanitaria San Carlos (IdISSC), Hospital Clínico San Carlos (HCSC), Madrid, Spain
[210], Universidad de Sevilla, Departamento de Enfermería, Seville, Spain
[211], Hospital General Universitario Gregorio Marañón (IiSGM), Madrid, Spain
[212], ERN-ITHACA-European Reference Network

[213], Unidad de Genética y Genómica Islas Baleares, Unidad de Diagnóstico Molecular y Genética Clínica, Hospital Universitario Son Espases, Islas Baleares, Spain
[214], Instituto de Investigación Sanitaria Islas Baleares (IdISBa), Islas Baleares, Spain
[215], Programa de Pós-Graduação em Biologia Animal, Universidade de Brasília, Brasília, Brazil
[216], Programa de Pós-Graduação em Ciências da Saúde, Universidade de Brasília, Brasília, Brazil
[217], Programa de Pós-Graduação Profissional em Ensino de Biologia, Universidade de Brasília, Brasília, Brazil
[218], Programa de Pós-Graduação em Ciências Médicas, Universidade de Brasília, Brasília, Brazil
[219], Anatomía Patológica, Instituto de Investigación Sanitaria San Carlos (IdISSC), Hospital Clínico San Carlos (HCSC), Madrid, Spain
[220], Tecnológico de Monterrey, Monterrey, Mexico
[221], Centro de Investigación en Anomalías Congénitas y Enfermedades Raras (CIACER), Universidad Icesi
[222], Departamento de Genetica, Fundación Valle del Lili
[223], Universidad Complutense de Madrid, Department of Immunology, Ophthalmology and ENT, Madrid, Spain
[224], Department of Neumology, Instituto de Investigación Sanitaria-Fundación Jiménez Díaz University Hospital - Universidad Autónoma de Madrid (IIS-FJD, UAM), Madrid, Spain
[225], Hospital Nuestra Señora de Sonsoles, Ávila, Spain
[226], Inditex, A Coruña, Spain
[227], Intensive Care Department, Instituto de Investigación Sanitaria-Fundación Jiménez Díaz University Hospital - Universidad Autónoma de Madrid (IIS-FJD, UAM), Madrid, Spain
[228], Hospital Universitario Príncipe de Asturias, Servicio de Microbiología Clínica, Madrid, Spain
[229], Universidad de Alcalá de Henares, Departamento de Biomedicina y Biotecnología, Facultad de Medicina y Ciencias de la Salud, Madrid, Spain
[230], GENYCA, Madrid, Spain
[231], Marinha do Brasil, Brazil
[232], Universidade de Brasília, Brasilia, Brazil
[233], Neuromuscular Diseases Unit, Department of Neurology, Hospital de la Santa Creu i Sant Pau, Universitat Autònoma de Barcelona, Barcelona, Spain
[234], Instituto Mexicano del Seguro Social (IMSS), Centro Médico Nacional Siglo XXI, Unidad de Investigación Médica en Enfermedades Infecciosas y Parasitarias, Mexico City, Mexico
[235], Catalan Institution of Research and Advanced Studies (ICREA), Barcelona, Spain
[236], Drug Research Centre, Institut d'Investigació Biomèdica Sant Pau, IIB-Sant Pau, Barcelona, Spain
[237], Departamento de Genetica, Clinica imbanaco
[238], Unidad de Cuidados Intensivos, Hospital Clínico Universitario de Santiago (CHUS), Sistema Galego de Saúde (SERGAS), Santiago de Compostela, Spain
[239], Department of Immunology, Hospital Universitario de Gran Canaria Dr. Negrín, Las Palmas de Gran Canaria, Spain
[240], Department of Clinical Sciences, University Fernando Pessoa Canarias, Las Palmas de Gran Canaria, Spain
[241], University Hospital Germans Trias i Pujol, Pediatrics Department, Badalona, Spain
[242], Department of Pathology, Biobank, Instituto de Investigación Sanitaria-Fundación Jiménez Díaz University Hospital - Universidad Autónoma de Madrid (IIS-FJD, UAM), Madrid, Spain
[243], Faculdade de Ciências da Saúde, Universidade de Brasília, Brasilia, Brazil
[244], Hospital Universitario Virgen de las Nieves, Servicio de Medicina Interna, Granada, Spain
[245], Fundación Universitaria de Ciencias de la Salud, Grupo de Ciencias Básicas en Salud (CBS), Bogotá, Colombia
[246], Sociedad de Cirugía de Bogotá, Hospital de San José, Bogotá, Colombia
[247], Universidad de Granada, Departamento Bioquímica, Biología Molecular e Inmunología III, Granada, Spain

[248], Hospital Infanta Elena, Allergy Unit, Valdemoro, Madrid, Spain
[249], Faculty of Medicine, Universidad Francisco de Vitoria, Madrid, Spain
[250], Hospital Universitario Infanta Leonor, Madrid, Spain
[251], Complutense University of Madrid, Madrid, Spain
[252], Gregorio Marañón Health Research Institute (IiSGM), Madrid, Spain
[253], Reumathology Service, Instituto de Investigación Sanitaria-Fundación Jiménez Díaz University Hospital - Universidad Autónoma de Madrid (IIS-FJD, UAM), Madrid, Spain
[254], Biobank, Puerta de Hierro-Segovia de Arana Health Research Institute, Madrid, Spain
[255], Universidad Rey Juan Carlos, Madrid, Spain
[256], The John Walton Muscular Dystrophy Research Centre, Newcastle University and Newcastle Hospitals NHS Foundation Trust, Newcastle upon Tyne, UK.
[257], Neuromuscular Unit, Neuropediatrics Department, Institut de Recerca Sant Joan de Déu, Hospital Sant Joan de Déu, Spain
[258], Fundació Docència i Recerca Mutua Terrassa, Terrassa, Spain
[259], Casa de Saúde São Lucas, Natal, Brazil
[260], Hospital Rio Grande, Rio Grande do Norte, Natal, Brazil
[261], Intensive Care Unit, Hospital Universitario de Gran Canaria Dr. Negrín, Las Palmas de Gran Canaria, Spain
[262], Universidad Fernando Pessoa Canarias, Las Palmas de Gran Canaria, Spain
[263], Hospital Clinico Universitario de Valladolid, Servicio de Anestesiologia y Reanimación, Valladolid, Spain
[264], Hospital Clinico Universitario de Valladolid, Servicio de Hematologia y Hemoterapia, Valladolid, Spain
[265], Hospital Universitario Lauro Wanderley, Brazil
[266], Hospital Universitario Infanta Leonor, Servicio de Medicina Interna, Madrid, Spain
[267], University Hospital of Burgos, Burgos, Spain
[268], Universidad de Sevilla, Seville, Spain
[269], Fundación Santa Fe de Bogota, Instituto de servicios medicos de Emergencia y trauma, Bogotá, Colombia
[270], Universidad de los Andes, Bogotá, Colombia
[271], Quironprevención, A Coruña, Spain
[272], Junta de Castilla y León, Consejería de Sanidad, Valladolid, Spain
[273], Gerencia Atención Primaria de Burgos, Burgos, Spain
[274], Immunogenetics-Histocompatibility group, Servicio de Inmunología, Instituto de Investigación Sanitaria Puerta de Hierro - Segovia de Arana, Madrid, Spain
[275], Hospital del Mar, Department of Infectious Diseases, Barcelona, Spain
[276], IMIM (Hospital del Mar Medical Research Institute, Institut Hospital del Mar d'Investigacions Mediques), Barcelona, Spain
[277], Universitat Autònoma de Barcelona, Department of Medicine, Spain
[278], Consejería de Sanidad, Comunidad de Madrid, Madrid, Spain
[279], Centro para el Desarrollo de la Investigación Científica, Asunción, Paraguay
[280], Genomics Division, Instituto Tecnológico y de Energías Renovables, Santa Cruz de Tenerife, Spain
